# Polygenic contributions to lithium augmentation outcomes in antidepressant non-responders with unipolar depression

**DOI:** 10.1101/2025.01.22.25320940

**Authors:** Julia Kraft, Pichit Buspavanich, Alice Braun, Georgia Panagiotaropolou, Peter Schlattmann, Hannah Buchbauer, Karl Heilbron, Urs Heilbronner, Thomas G. Schulze, Stephan Ripke, Roland Ricken, Mazda Adli

## Abstract

**Objective:** Lithium augmentation (LA) is an effective treatment for patients with major depression after inadequate antidepressant response, but therapeutic outcomes vary considerably between individuals. Molecular studies could yield novel insights into treatment prediction to enable personalized therapy choices. Here, we investigated the effects of polygenic risk scores (PRS) for schizophrenia (SCZ), major depressive disorder (MDD), and bipolar disorder (BIP) on clinical outcomes following LA.

**Methods:** Recent GWAS summary statistics were used to construct disorder-specific PRS in lithium-augmented MDD patients who participated in a prospective study after poor response to at least one antidepressant drug. Depressive symptoms were assessed for four weeks or longer using the Hamilton Depression Rating Scale (HAMD). Hazard ratios (HR) of favorable outcomes, response (≥ 50% reduction in HAMD composite scores) and remission (HAMD ≤ 7), were estimated by Cox proportional hazards regression models adjusted for ancestry, demographic, and clinical covariates.

**Results:** In 193 patients, BIP-PRS was positively associated with both response (HR = 1.29, 95% CI = 1.02-1.63, p = 0.03, Nagelkerke R^2^ = 2.51%) and remission (HR = 1.52, 95% CI = 1.14-2.04, p = 0.004, Nagelkerke R^2^ = 4.53%) after LA. Our data further suggest that individuals who carry a lower polygenic burden for MDD tend to respond better to LA (HR = 0.81, 95% CI = 0.66-1.00, p = 0.048, Nagelkerke R^2^ = 1.99%). No associations were observed between SCZ-PRS and either clinical outcome (p > 0.05).

**Conclusions:** Our findings indicate that individuals at higher polygenic risk for BIP and lower polygenic risk for MDD are more likely to benefit from augmentation with lithium. If replicated, PRS may inform future efforts to establish clinical prediction models for LA outcomes in unipolar depression.

## 1 Introduction

Major depressive disorder (MDD) is a prevalent psychiatric condition primarily characterized by depressive mood and anhedonia. The use of an antidepressant drug (AD) can reduce some of the symptoms; however, only ∼37% of patients treated for MDD fully remit after one adequate AD trial (1). One effective approach for the treatment and management of patients without complete antidepressant response is pharmacological augmentation of ADs.

Lithium, a first-line mood stabilizer in the treatment of bipolar disorder (BIP), is a well-established augmenting agent, and its adjunctive administration after antidepressant non-response and in treatment-resistant depression (TRD). has been recommended by most therapy guidelines (2). Despite its demonstrated efficacy in combination with both first- and second-generation AD (3), as well as its putative value in lowering suicide risk and long-term prevention of recurring depressive episodes (4), potential side effects of lithium may cause reluctance to initiate lithium augmentation (LA) in practice. Patients may experience weight gain, hypothyroidism, and kidney impairment in long-term use of lithium (5), especially at higher dosages. To prevent severe lithium intoxication monitoring of drug plasma levels and blood markers is required. At present, validated biomarkers that can guide clinical decision-making on which patients might benefit from LA are lacking.

Prior research indicates that variability in response to ADs and lithium is influenced by genetic factors. Common genetic variation was estimated to explain up to 30% of the variation in lithium response among patients with BIP (6) and, similarly, 20-40% of the AD response variability in individuals with MDD (7), highlighting the polygenic nature of both traits. Due to modest sample sizes, genome-wide association studies (GWAS) have had limited success in robustly identifying variant-level associations for AD response (7) and lithium response (6,8). Aggregated genetic measures such as polygenic risk scores (PRS) may be more useful for predicting treatment outcomes.

In essence, a PRS reflects an individual’s propensity towards a trait or disease that is attributable to effects of single nucleotide polymorphisms (SNPs) across the genome. Previous studies suggest that MDD patients with lower responses to AD treatment tend to carry a higher polygenic burden for MDD (9,10) and schizophrenia (SCZ) (7,11). Consistent with this observation, PRSs for MDD and SCZ were also negatively associated with lithium response in individuals with BIP (12,13). While a predisposition for BIP has been suspected to affect AD responsiveness, a direct genetic link has yet to be established (10,14). It remains unclear whether these findings extend to therapeutic effects of LA in unipolar depression. Candidate gene studies reported positive results for LA-associated side effects (15,16) and outcomes (17,18), nevertheless, single SNPs may not be sufficient to capture much of the genetic signal contributing to individual differences in response given the evidence supporting polygenic drug response models.

Here, we aim to assess the influence of polygenic risk for three major psychiatric disorders (SCZ, BIP, MDD) on treatment outcomes in a prospective cohort of MDD patients undergoing LA following inadequate AD response. The goal of this study is to generate new insights into potential pharmacogenetic factors underlying LA to facilitate personalized therapy strategies in MDD patients with refractory symptoms.

## 2 Methods

### 2.1 Study population

A total of 332 patients were recruited in a prospective cohort study to identify predictors of LA treatment outcomes in MDD. Recruitment was carried out between 2008 and 2020 at 13 psychiatric facilities within the Berlin Research Network on Depression, Berlin, Germany. All participants a) met diagnostic criteria for unipolar depression; b) were at least 18 years old; c) did not adequately respond to at least one prior antidepressant monotherapy trial; d) had a clinical indication for LA. Diagnosis was confirmed by the Mini-International Neuropsychiatric Interview. Exclusion criteria were contraindication for lithium treatment (e.g., severe kidney insufficiency), depressive symptoms due to other organic or psychiatric disorders, dementia or other severe neurological disorders, substance dependency within the last 6 months, and antisocial personality disorder.

After excluding participants without quality-controlled genetic data (n=37) and complete phenotype data (n=9) 286 individuals remained for analysis in the broadly defined study population. We further excluded individuals with a treatment duration of less than 28 days, diagnostic switches during or after LA, a baseline HAMD-17 score below 12, changes in co-medication during LA, as well as pre-treatment that could potentially influence clinical outcomes. This resulted in a narrowly defined study population of 193 participants. The sample selection procedure is illustrated in **Figure S1**.

### 2.2 Study proceedings and outcome measures

Study enrollment was preceded by a screening phase during which non-response to antidepressant treatment (administered at a sufficient dose for ≥ 4 weeks) and eligibility were determined. Before LA was initiated, patients underwent a baseline assessment including diagnostic interviews, current symptom severity, and collection of a blood sample for genotyping. During augmentation, depressive symptoms and serum lithium concentrations were measured over a period of at least 4 weeks. Study procedures continued until therapeutic lithium levels (≥ 0.5 mmol/L) were observed for ≥ 2 weeks. Symptom severity was evaluated weekly by administration of the 17-item Hamilton Depression Rating Scale (HAMD-17). Following standard conventions, remission was defined as a HAMD-17 score ≤ 7 and response as a ≥ 50% reduction in HAMD-17 score. All patients provided written informed consent to participate in the study. The study protocol was reviewed and approved by the institutional ethics board.

### 2.3 Genotyping

Genomic DNA was extracted from frozen EDTA whole-blood samples using established standard protocols. The quality and quantity of DNA samples were verified by gel electrophoresis, UV-vis spectroscopy (NanoDrop; Thermo Scientific, Thermo Fisher Scientific, Inc., MA, USA) and a fluorescence-based dsDNA quantification assay (PicoGreen; Invitrogen, Thermo Fisher Scientific, Inc., MA, USA). All samples were then assayed on the Illumina Infinium Global Screening Array BeadChip (Illumina, San Diego, CA) version 3.0 covering more than 700,000 SNPs.

### 2.4 Genetic data

#### 2.4.1 Quality checks and imputation

The RICOPILI pipeline was used to process genome-wide genotype data (19). Quality checks (QC) were applied to exclude SNPs with low genotyping rates (< 0.98), Hardy-Weinberg equilibrium violations (P > 10^-6^), autosomal heterozygosity deviations (| Fhet | > 0.2), as well as samples with low call rates (< 0.98) and sex inconsistencies (discrepancies between genotype and reported sex). A total of 316 individuals and 544,626 SNPs passed these QC criteria.

After technical QC, relatedness testing was performed on 91,320 LD-pruned SNPs (R^2^ > 0.02) using PLINK v1.9 (20). One related pair (PI-HAT > 0.2) was identified in the sample, and one of these individuals was excluded at random. Principal component analysis (PCA) was performed on the same set of SNPs, and the first two principal components (PCs) were plotted to assess ancestral heterogeneity. Based on visual clustering we excluded 20 individuals to retain only European participants.

Genome-wide SNP data of all remaining individuals were then pre-phased with EAGLE version 2.4.1 (21) and imputed with MINIMAC3 (22) using the Haplotype Reference Consortium (HRC) release 1.1 as the reference panel.

#### 2.4.2 Polygenic scoring

Summary statistics from well-powered GWAS for MDD (23), SCZ (24), BIP (25) were selected as training datasets (for details see **Table S1**). We subset these summary statistics to SNPs that intersected with the imputation reference panel (HRC), had a minor allele frequency (MAF) > 0.05, and had imputation quality (INFO score) > 0.9. When appropriate, we used summary statistics excluding Berlin-based cohorts to avoid sample overlap. Posterior SNP effect sizes were then derived using continuous shrinkage priors implemented in PRS-CS (26) with default settings (--a=1 --b=0.5 --n_iter=1000 --n_burnin=500 --thin=5) and a linkage disequilibrium (LD) matrix constructed using the European 1000 Genomes (phase 3) reference panel. To obtain a single PRS for each disorder in the target sample, imputed dosages of the effect alleles were multiplied by corresponding posterior SNP effect sizes (log odds ratio [OR]) across the genome and summed over each individual. PRSs were computed in PLINK v1.9 (20) for MDD (745,573 SNPs), SCZ (983,271 SNPs), and BIP (886,211 SNPs), respectively. For comparison, we also generated PRS at ten different p thresholds (PTs) ranging between PT < 5 x 10^-8^ and PT < 1 (no filter) using clumped summary statistics (R^2^ > 0.1 within a 500kb window). The number of SNPs retained per trait at each PT is summarized in **Table S1**. To improve interpretability, all PRS were scaled to have a mean of zero and standard deviation (SD) of one.

### 2.5 Statistical analysis

Cox proportional hazards models were used to investigate time-to-event relationships between PRSs for MDD, BIP, SCZ and clinical outcomes of LA (response and remission) in the narrowly defined sample. The full model included the following covariates: age, sex, HAMD-17 scores at baseline, sufficient lithium levels, the first three ancestry components (PC 1-3) and PC 19-20 (associated with either outcome). Adjusted hazard ratios (HRs) for favorable treatment outcomes with 95% confidence intervals (CI) were calculated for each PRS individually. The amount of variability in LA response and remission attributable to polygenic variation was quantified using Nagelkerke R^2^. Harrell’s C-index was reported as a measure of concordance to assess overall model performance.

Individuals were then stratified into tertiles based on their polygenic loading for each trait. Treatment outcomes were summarized within each PRS stratum (percentage of responders and remitters). Associations between both outcomes and each PRS stratum were then assessed in multivariable Cox regression models using the bottom PRS stratum as the reference group. Unadjusted and adjusted HRs including 95% confidence intervals (CIs) were reported for each PRS stratum using the same covariates as described above. Cumulative incidence curves were shown to visualize time-dependent differences in treatment outcomes for low (1st tertile), average, and high (3rd tertile) PRS groups.

To evaluate the robustness of the polygenic associations we conducted several sensitivity analyses. First, we repeated all analyses, including stratification in the broad study sample. Second, we examined polygenic associations in both study populations when all PRS were jointly modeled (multi-PRS model). Third, we estimated associations between each PRS and clinical outcomes in the narrow and broad cohort using PRSs obtained from an LD-based clumping and p thresholding approach (C+PT) across multiple thresholds.

Two-sided p-values are reported, and a significance level of p < 0.05 was applied for all tests. R software (R version 4.0.3 (2020-10-10)) was used for statistical analysis.

## 3 Results

### 3.1 Demographic and clinical characteristics

Out of 332 participants enrolled in the study, 286 individuals remained in the broad study sample which includes 193 individuals augmented according to narrow study criteria (**Figure S1**, **Table 1**). In the narrowly defined sample, the average age at study entry was 49.45 (SD=13.41) years and the proportion of females was 61%. The response and remission rates in the narrow sample were 47% and 32%, respectively. Most patients (77%) received either an SSRI or SNRI when LA was initiated (**Table S2**). The sociodemographic and clinical characteristics of the broad sample are summarized in **Table 1**.

**Table 1.**
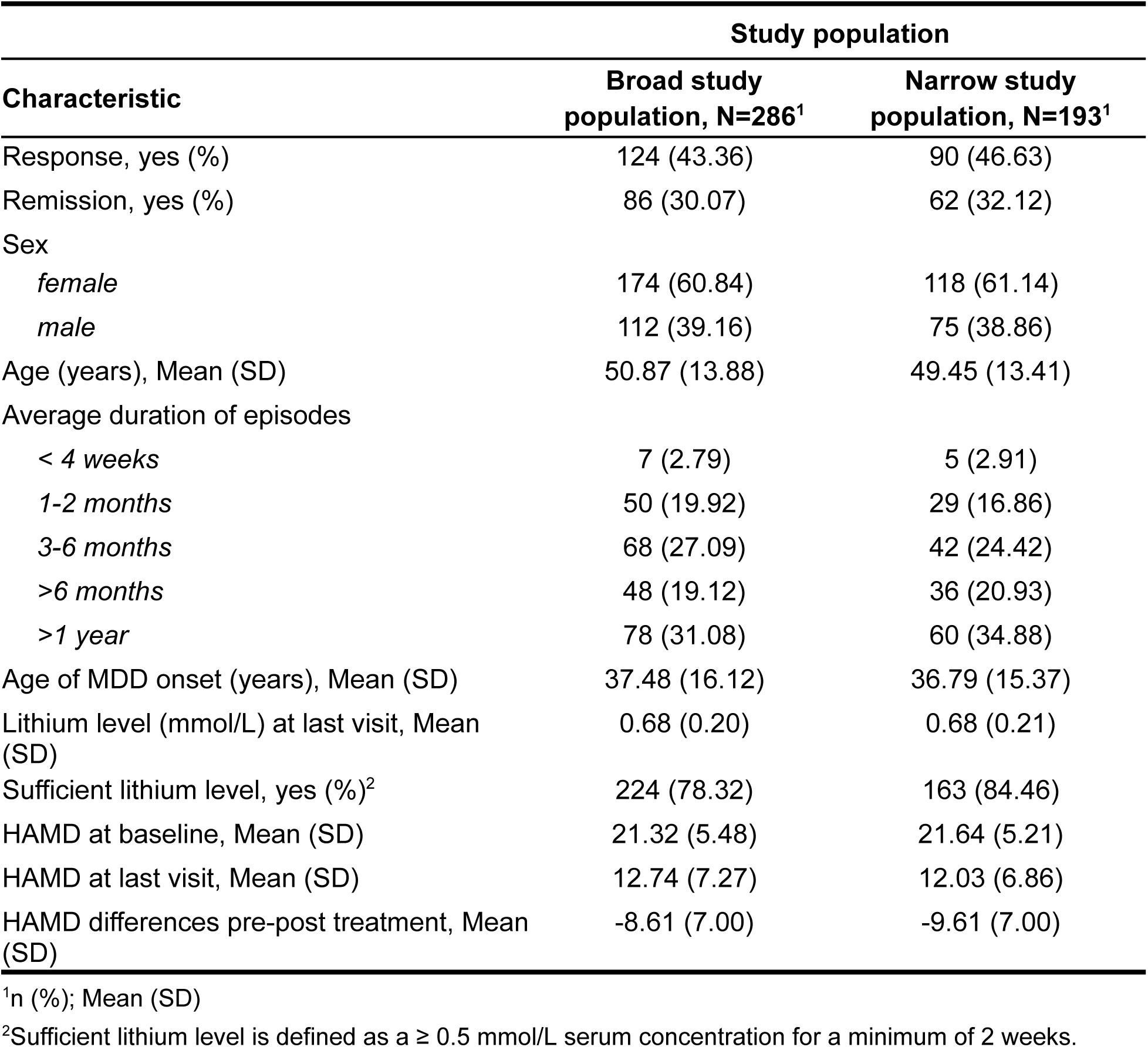
Summary of demographic and clinical data.

| Characteristic | Study population |  |
| --- | --- | --- |
|  | Broad study population, N=286<sup>1</sup> | Narrow study population, N=193<sup>1</sup> |
| Response, yes (%) | 124 (43.36) | 90 (46.63) |
| Remission, yes (%) | 86 (30.07) | 62 (32.12) |
| Sex |  |  |
| *female* | 174 (60.84) | 118 (61.14) |
| *male* | 112 (39.16) | 75 (38.86) |
| Age (years), Mean (SD) | 50.87 (13.88) | 49.45 (13.41) |
| Average duration of episodes |  |  |
| < 4 weeks | 7 (2.79) | 5 (2.91) |
| 1-2 months | 50 (19.92) | 29 (16.86) |
| 3-6 months | 68 (27.09) | 42 (24.42) |
| >6 months | 48 (19.12) | 36 (20.93) |
| >1 year | 78 (31.08) | 60 (34.88) |
| Age of MDD onset (years), Mean (SD) | 37.48 (16.12) | 36.79 (15.37) |
| Lithium level (mmol/L) at last visit, Mean (SD) | 0.68 (0.20) | 0.68 (0.21) |
| Sufficient lithium level, yes (%)<sup>2</sup> | 224 (78.32) | 163 (84.46) |
| HAMD at baseline, Mean (SD) | 21.32 (5.48) | 21.64 (5.21) |
| HAMD at last visit, Mean (SD) | 12.74 (7.27) | 12.03 (6.86) |
| HAMD differences pre-post treatment, Mean (SD) | -8.61 (7.00) | -9.61 (7.00) |

<sup>1</sup>n (%); Mean (SD)<sup>2</sup>Sufficient lithium level is defined as a $\geq 0.5$ mmol/L serum concentration for a minimum of 2 weeks.

### 3.2 Polygenic associations with LA outcomes

Time-dependent relationships between binary treatment outcomes and PRSs for SCZ, BIP and MDD were estimated in independent Cox regression models (**Figure 1** and **Table S3**). In the narrowly defined study population, the BIP-PRS was positively associated with response to LA (HR = 1.29, 95% CI = 1.02-1.63, p = 0.03, C = 0.62) and remission (HR = 1.52, 95% CI = 1.14-2.04, p = 0.004, C = 0.72), explaining 2.5% and 4.5% of the variability in response and remission, respectively. When patients were stratified into tertiles according to BIP-PRS, the response rate was 21% higher in the highest BIP-PRS tertile (56% responders) relative to the lowest tertile (35% responders). Patients in the highest BIP-PRS tertile had an adjusted hazard ratio (HR) of 2.02 (95% CI = 1.15-3.53) for response and 2.26 (95% CI = 1.17-4.36) for remission in comparison to the lowest tertile (**Table 2** and **Figure S2**).

**Figure 1.**
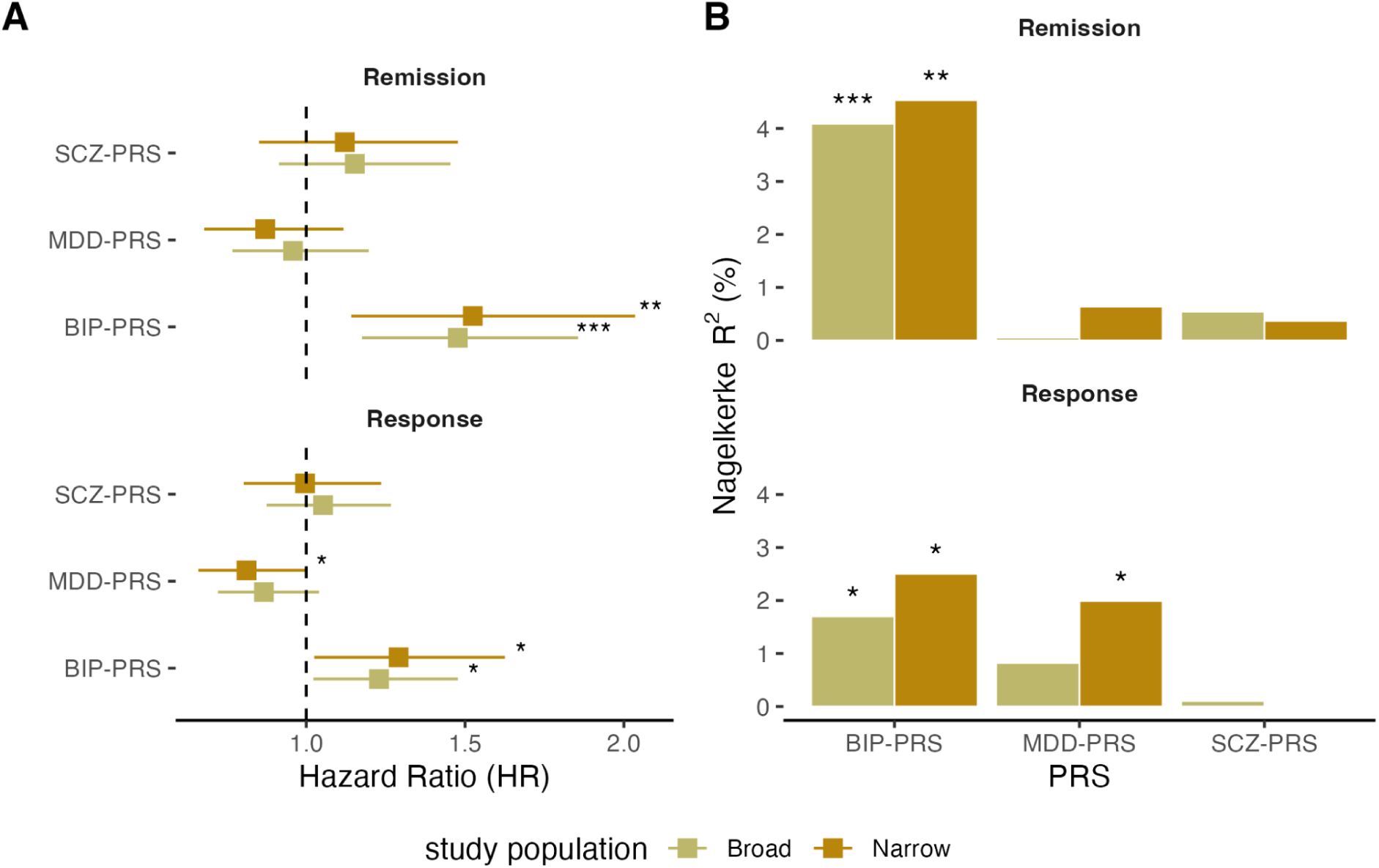
Associations between polygenic liability for schizophrenia (SCZ), major depressive disorder (MDD) and bipolar disorder (BIP) and clinical outcomes of LA (response and remission) among participants who met narrow study criteria and in the broadly defined study population. A) Hazard ratios (HR) showing the effect of a one SD increase in PRSs on remission and response to LA, HR >1 indicate a positive relationship between favorable outcomes and increasing polygenic risk. Error bars represent 95% confidence intervals. All HRs are adjusted for age, sex, baseline HAMD-17 score, sufficient lithium level and five principal components (PCs). B) Proportion of variability in remission and response to LA explained by each PRS (Nagelkerke R^2^). Annotated p-values: * p < 0.05; ** p < 0.01; *** p < 0.001.

**Table 2.**
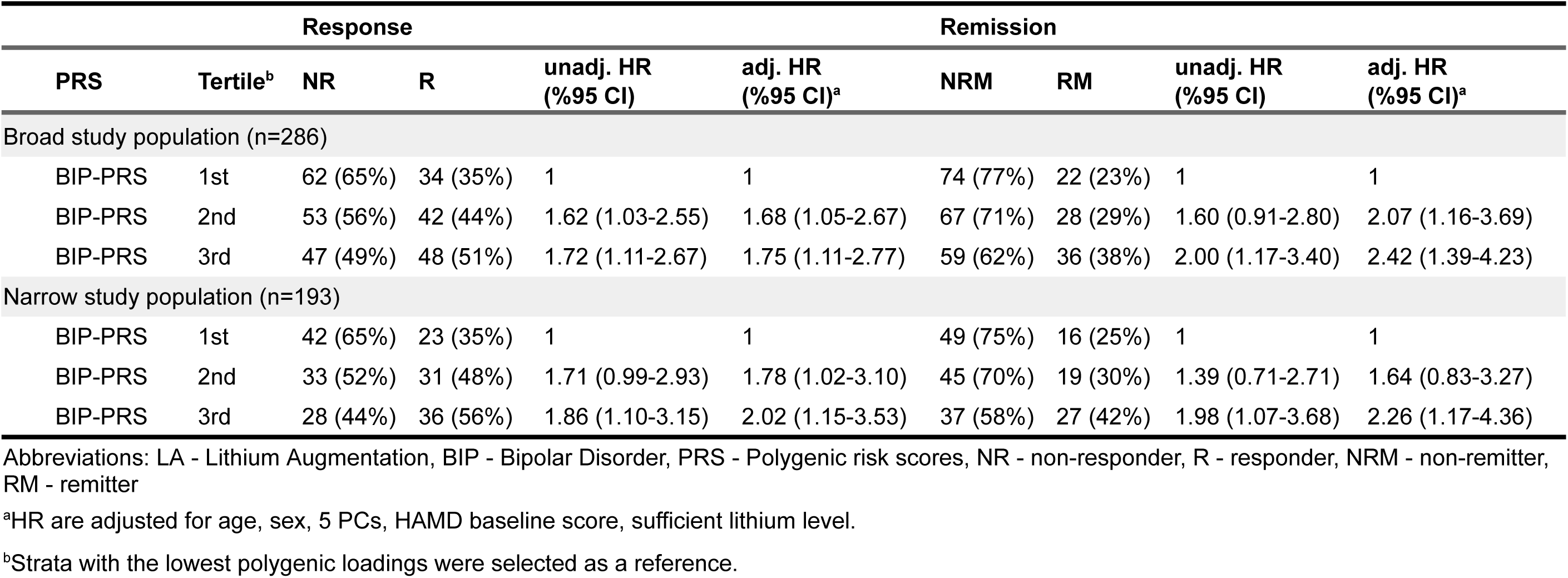
Hazard ratios (HR) for favorable LA outcomes by BIP-PRS strata.

We found a weak negative relationship between MDD-PRS and LA outcomes indicating that response to LA is associated with a lower MDD-PRS (HR = 0.81, 95% CI = 0.66-1.00, p = 0.048, C = 0.61, Nagelkerke R^2^ = 1.99%). The proportion of responders in the lowest MDD-PRS tertile was larger (48%) compared to the highest MDD-PRS tertile (39%, see **Table S4**). While LA remitters also tended to carry a lower polygenic burden for MDD, this association did not reach statistical significance (HR = 0.87, 95% CI = 0.68-1.12, p = 0.276, C = 0.71).

Finally, no statistically significant associations were observed between SCZ-PRS and response (HR = 1.00, 95% CI = 0.80-1.24, p = 0.972, C = 0.59) or remission (HR = 1.12, 95% CI = 0.85-1.48, p = 0.416, C = 0.71).

### 3.3 Sensitivity analyses

The primary association analyses were repeated in the broadly defined study population and yielded similar results with respect to BIP-PRS and SCZ-PRS (**Figure 1**, **Table S3**, and **Table 2**). While MDD-PRS was not significantly associated with response, the direction of effect is consistent with the primary analysis (HR = 0.87, 95% CI = 0.72-1.04, p = 0.124).

Because the PRS for mood disorders and schizophrenia were correlated (**Figure S3**), we next investigated associations between clinical outcomes and each PRS in a combined Cox regression model (see **Table S5**, **Figure S4**). While SCZ-PRS showed no effect on either response (HR = 0.91, 95% CI = 0.70-1.17, p = 0.451) or remission (HR = 0.97, 95% CI = 0.70-1.34, p = 0.843), BIP-PRS remained associated with response to LA (HR = 1.43, 95% CI = 1.10-1.86, p = 0.007) as well as remission (HR = 1.60, 95% CI = 1.17-2.21, p = 0.004) in the narrow study population. In the multi-PRS model, MDD-PRS was also significantly associated with a poor LA response (HR = 0.77, 95% CI = 0.62-0.96, p = 0.022) but not remission (HR = 0.82, 95% CI = 0.63-1.06, p = 0.131). Notably, predictive performance of the multi-PRS model was slightly higher than that of any single-PRS model in terms of response (2.1-5.9% improvement in concordance), whereas jointly modeling PRS did not improve the performance in predicting remission (see **Figure S5**)

As a third sensitivity analysis, PRS obtained from a standard approach (clumping and p-value thresholding [C+PT]) were used to estimate polygenic associations with both clinical outcomes (see **Table S6** and **Figure S6**). In line with our primary results in the narrow population, response to LA was predicted by a higher PRS for BIP (HR = 1.41, 95% CI = 1.13-1.76, p = 0.003 at PT<0.05) and a lower PRS for MDD (HR = 0.77, 95% CI = 0.62-0.97, p = 0.026 at PT < 0.1) when a C+PT polygenic scoring method was used. We also observed an association between BIP-PRS and remission after LA (HR = 1.52, 95% CI = 1.17-1.96, p = 0.002 at PT < 0.5).

## 4 Discussion

To the best of our knowledge, this is the first study to examine relationships between genetic risk for major psychiatric disorders and clinical outcomes related to depressive symptom severity after LA in patients with antidepressant non-response. We demonstrated that individuals with a higher polygenic burden for bipolar disorder respond better to LA and are more likely to achieve remission at the study endpoint, irrespective of their polygenic liability for major depression and schizophrenia. Sensitivity analyses further underline the robustness of our findings across varying sample compositions and polygenic scoring methods.

Notably, lithium response in patients with bipolar disorder has not previously been linked to an increased polygenic burden for BIP (27). In patients receiving LA higher response rates were observed among individuals with a final diagnosis of bipolar depression (28), family history of affective disorders (28) or bipolar disorder (29) as well as for individuals who later converted to bipolar depression (29). Thus, the hypothesis emerged that LA responders could have covert or subthreshold bipolar depression (30). However, studies supporting this claim rely on very small, selected samples (N < 80) and should therefore be interpreted with caution. Our findings also raise the possibility that the predictive value of BIP-PRS may be specific to unipolar depression. Here, potential misdiagnoses were minimized by screening for manic episodes at baseline and excluding patients with diagnostic shifts after chart review. Nevertheless, longer follow-up periods are needed to disentangle polygenic contributions to therapeutic effects and diagnostic progression, especially conversion to bipolar depression.

In line with previously-reported associations between MDD polygenic risk and lithium response in BIP (12,31), we found evidence that LA responders with an MDD diagnosis are also more likely to carry a lower polygenic burden for MDD. In patients with MDD, non-responders to AD tended to show higher MDD-PRS (10), which may also modulate antidepressant effects to a concomitant AD during LA. The exact mechanism of action remains unclear, but one possibility is that lithium augments antidepressant response by synergistic activation of neurotransmitter and signaling pathways relevant to mood regulation (32). Further dissection of genetic signals at pathway level may lead to an improved understanding of the neurobiological processes behind AD and lithium response.

Unlike prior research on bipolar disorder, which indicated poor lithium response in patients with higher SCZ-PRS (13), we did not observe an association between SCZ-PRS and clinical outcomes after LA in unipolar depression. However, the effect of SCZ-PRS on lithium response in BIP appears to be largely mediated by treatment adherence, that is a higher genetic load for SCZ is associated with worse adherence resulting in an overall reduced response to lithium (31). Low rates of adherence to mood stabilizers are a well-known issue in clinical practice (33), particularly among patients with comorbidities like substance abuse (34), and impact clinical outcomes negatively (35). In this study setting, we controlled for suboptimal drug exposure using longitudinal data on serum lithium levels and excluded patients with substance dependency, which could potentially account for conflicting results.

Interestingly, our results align with the largest study to date examining influences of PRS for major psychiatric disorders on ECT effectiveness in depression (36). In this study improvement after ECT was positively associated with BIP-PRS and negatively associated with MDD-PRS (36). While it is premature to draw any conclusions, past and current findings support the notion of an MDD subtype that is less responsive to non-pharmacological interventions like ECT (36) and pharmacotherapy (10), yet at greater risk of unfavorable outcomes, such as recurrent depressive episodes (37) or suicide attempts (38).

Our results must be considered in light of several limitations. On the basis of previously published studies in mood disorders, PRS effects on treatment outcomes are expected to be small or moderate at best. Although this cohort is clinically well-characterized, the study sample size remains a limiting factor, and our analysis may be underpowered to detect small polygenic effects.

Second, we did not apply a correction for multiple testing, as our primary analysis included only six statistical tests. If we had applied a Bonferroni correction (alpha = 0.05/3 = 0.017) for each outcome, the association between BIP-PRS and remission would have remained statistically significant.

Additionally, all findings need to be replicated in independent and, preferably, larger samples to assess their generalizability beyond this cohort, especially to populations of non-European ancestry. As the focus of this study was on improvements in depressive symptoms during the first weeks after LA initiation, uncertainties exist regarding long-term outcomes. Differences in outcome definitions may also offer at least a partial explanation for inconsistent findings in lithium-treated BIP cases, that are retrospectively assessed for their overall response to long-term treatment using a composite measure (ALDA scale).

Despite inconclusive evidence on clinical predictors of LA (39), combining clinical and genetic data may still result in improved classification of treatment outcomes as recently demonstrated by works in BIP (40). Given the modest explanatory power of single PRS, such studies are an important first step toward developing prognostic algorithms for personalized treatment strategies, not only for first-line but also second-line therapies.

While our study enhances the understanding of the genetic factors influencing treatment outcomes after lithium augmentation (LA) in unipolar depression, further research is needed to validate our findings and explore additional genetic and non-genetic factors that may affect outcomes. Identifying genetic markers linked to treatment responses could improve clinical practice by enabling clinicians to personalize treatment strategies based on patients’ genetic profiles. Such personalized approaches could enhance treatment efficacy, minimize adverse effects, and optimize patient outcomes. In addition, integrating genetic data with clinical and demographic information may support the development of prognostic algorithms to guide treatment decisions.

## Data Availability

Summary statistics of the schizophrenia, major depressive disorder, and bipolar disorder GWAS meta-analyses are freely available for download on the website of the Psychiatric Genomics Consortium (PGC): https://pgc.unc.edu/for-researchers/download-results/

## Acknowledgements

We would like to thank the members of the Berlin Research Network on Depression (https://www.berliner-wissenschaftsnetz-depression.de/). The recruitment of patients who generously volunteered to participate in this project would not have been possible without the invaluable support of the many colleagues from this network, encompassing both academic and non-academic institutions across the greater Berlin region. We further thank the Major Depressive Disorder, Schizophrenia, and Bipolar Disorder working groups of the Psychiatric Genomics Consortium (PGC) for providing summary statistics that were used as a training dataset. We would like to thank the research participants and employees of 23andMe, Inc. for making this work possible.

