## Supplementary Material for "Polygenic contributions to lithium augmentation outcomes in antidepressant non-responders with unipolar depression"

**Figure S1.** Flow chart illustrating sample selection and exclusion criteria.

**Figure S2.** Cumulative incidence curves for low, average, and high polygenic risk groups based on MDD-, BIP-, and SCZ-PRS.

**Figure S3.** Relationships between polygenic risk scores for SCZ, BIP, MDD.

**Figure S4.** Association between polygenic risk scores for SCZ, BIP, MDD and LA outcomes in joint cox regression models (multi-PRS model).

**Figure S5.** Predictive performance of single- and multi-PRS models.

**Figure S6** Association between polygenic risk scores for SCZ, BIP, MDD and LA outcomes (C+PT).

**Table S1.** Number of SNPs included in polygenic risk scores.

**Table S2.** Summary table of antidepressants and psychotropic co-medication at baseline.

**Table S3.** Association between polygenic risk scores for SCZ, BIP, MDD and clinical outcomes after LA.

**Table S4.** Hazard ratios (HR) for favorable LA outcomes by PRS strata.

**Table S5.** Association between polygenic risk scores for SCZ, BIP, MDD and clinical outcomes after LA (multi-PRS model).

**Table S6.** Association between polygenic risk scores for SCZ, BIP, MDD at ten p-value thresholds (C+PT) and clinical outcomes. after LA.

**Abbreviations**

SCZ - Schizophrenia

BIP - Bipolar Disorder

MDD - Major Depressive Disorder

LA - Lithium Augmentation

PC - Principal Component

PRS - Polygenic Risk Score

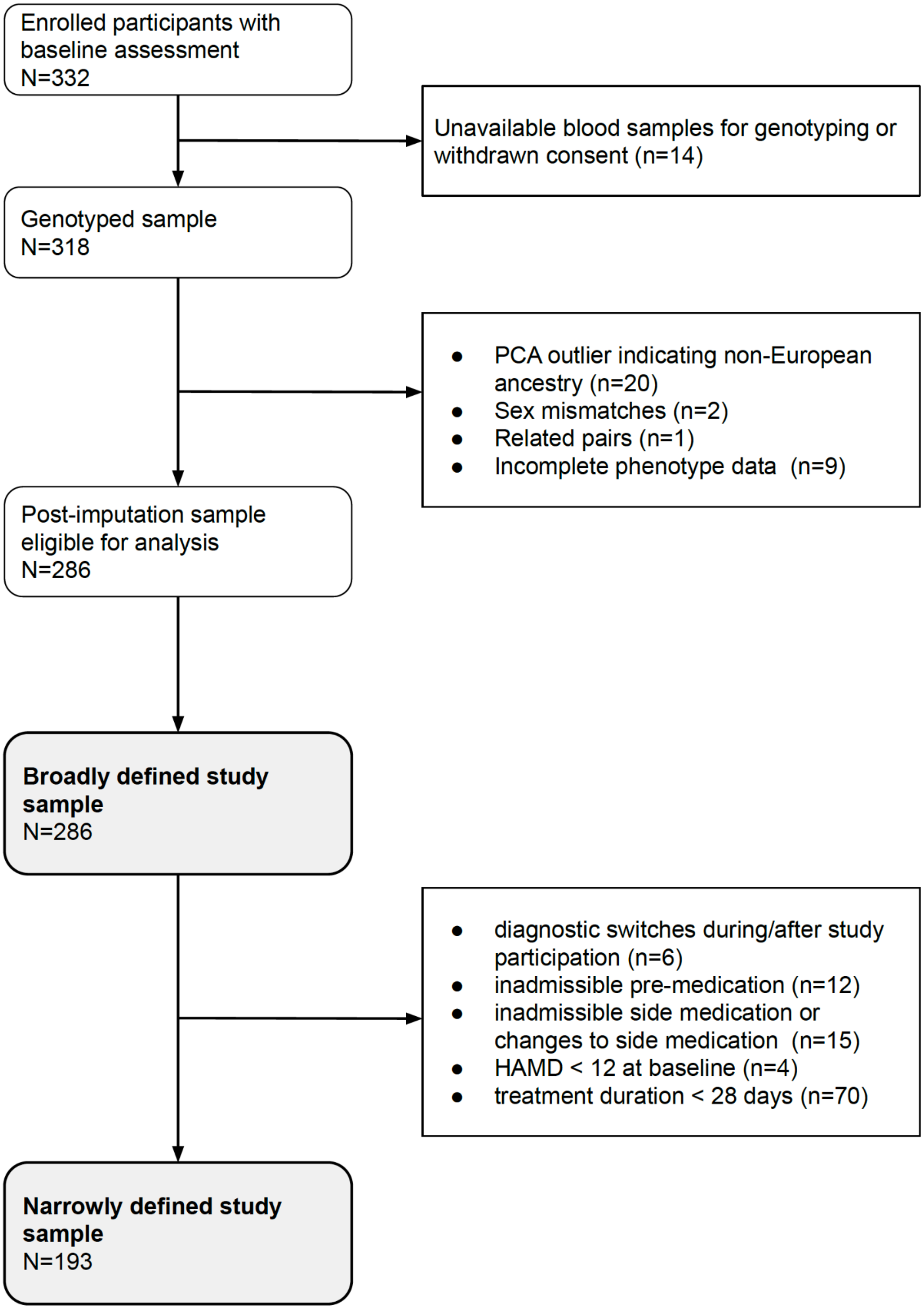


**Figure S1.** Flow chart illustrating sample selection and exclusion criteria.

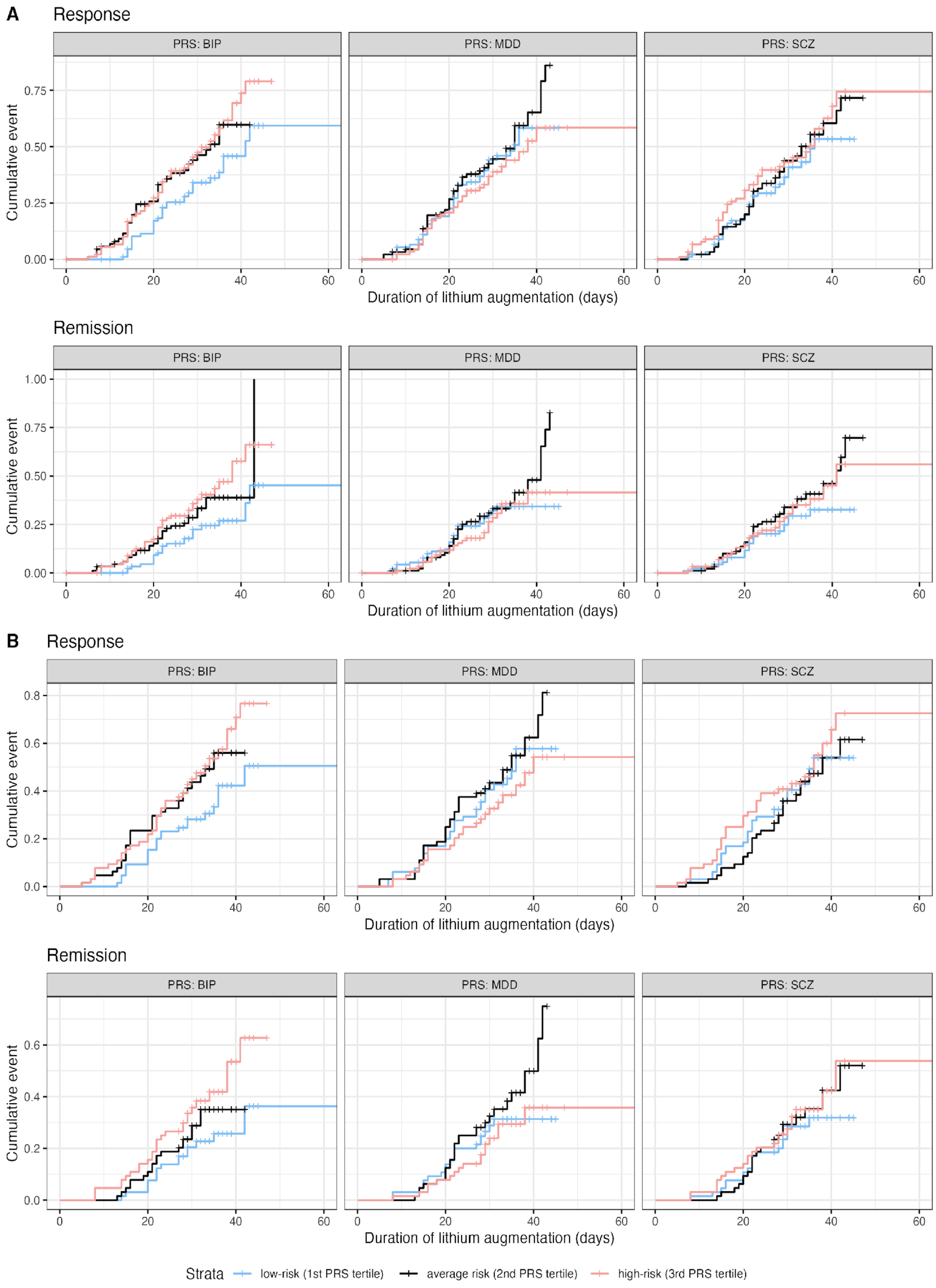


**Figure S2.** Cumulative incidence curves for low, average, and high polygenic risk groups based on MDD-, BIP-, and SCZ-PRS. Kaplan Meier cumulative event curves are shown for patients in low (light blue), average (dark blue) and high PRS (light pink) strata. Subfigure A) shows curves of the broadly defined study sample (n=286) and B) for the analysis conducted in the narrowly defined study sample (n=193). Time-to-event curves are shown for remission and response per PRS (horizontal panels).

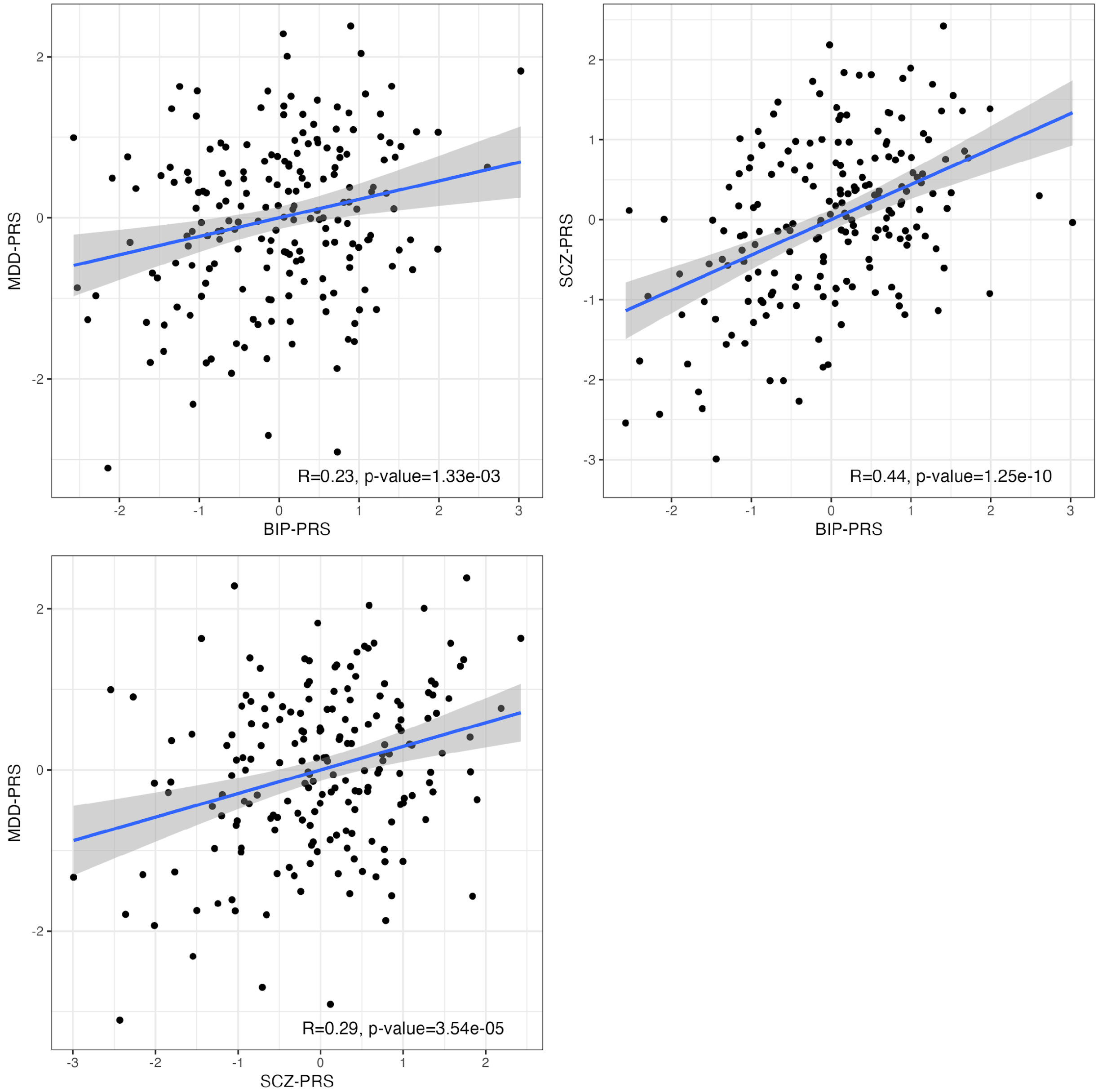


**Figure S3**. Relationships between polygenic risk scores for SCZ, BIP, MDD. Scatterplots displaying relationships between PRS for MDD, SCZ, and BIP in the narrow study population (n=193). Fitted lines are added to each plot (blue) assuming linear relationships between PRS, 95% confidence intervals are shaded grey. Pearson correlation coefficient (R) for each PRS pairing and p are shown at the bottom right corner.

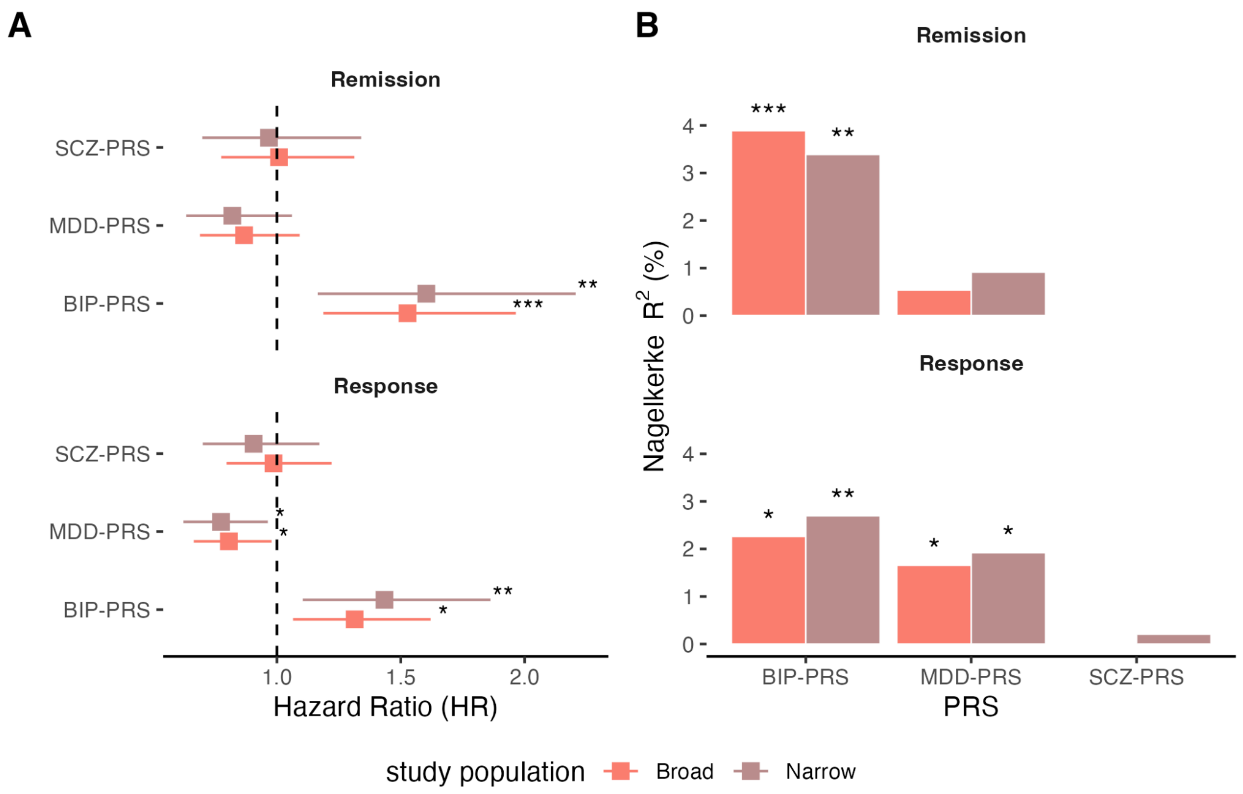


**Figure S4.** Association between polygenic risk scores for SCZ, BIP, MDD and LA outcomes in joint cox regression models (multi-PRS model). A) Forest plot showing HR (95% CI) of clinical outcomes after LA with PRS for three psychiatric disorders (SCZ, BIP, MDD) estimated in a joint cox regression model. HR above 1 indicate a favorable treatment outcome. B) Bar plot showing variation in outcome explained (Nagelkerke $R^2$) per PRS in both target study populations. Displayed p-values: * $p < 0.05$; ** $p < 0.01$; *** $p < 0.001$.

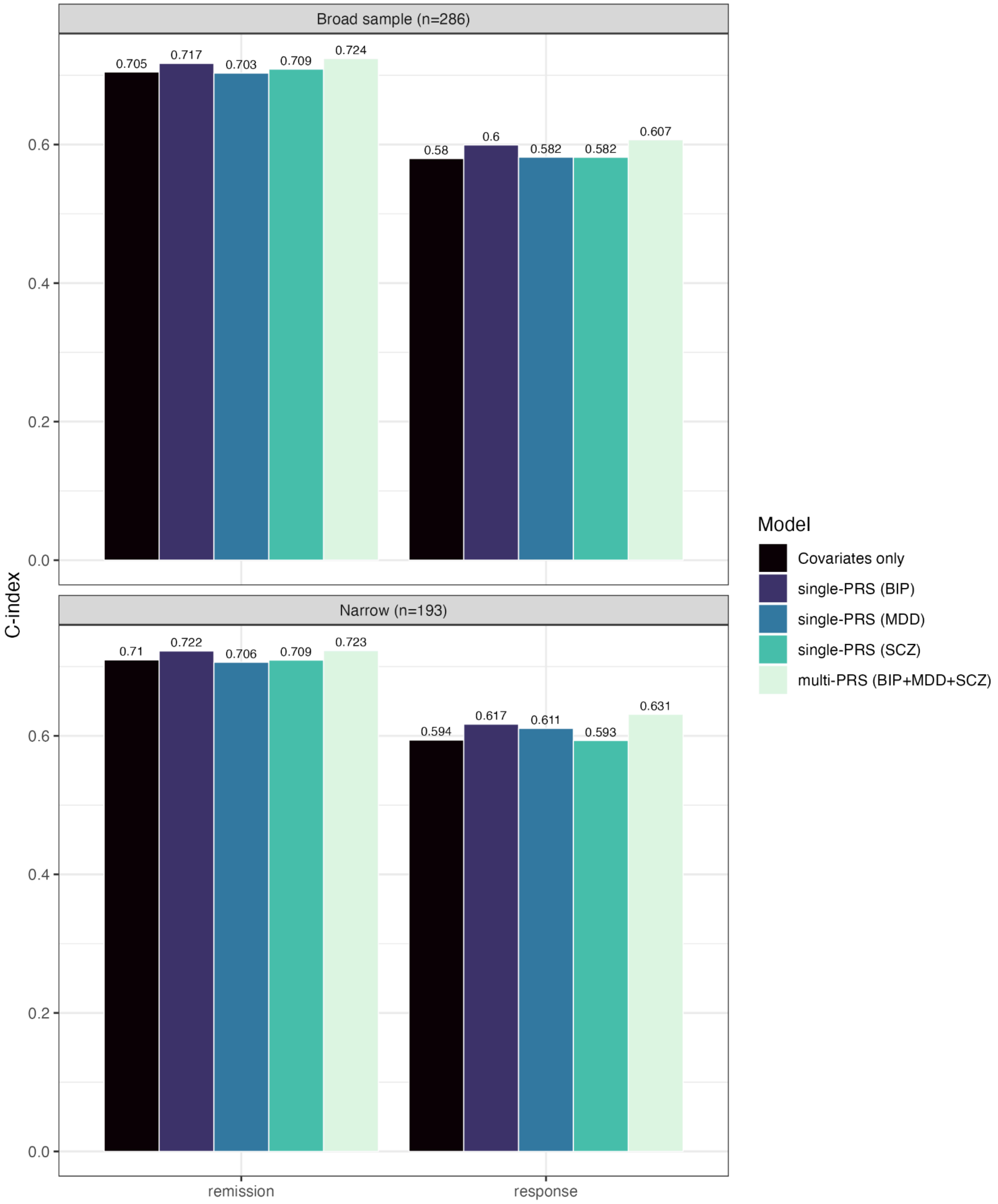


**Figure S5.** Predictive performance of single- and multi-PRS models. Bar plots displaying the predictive performance (C-index) of single- and multi-PRS models including covariates (PCs, sufficient lithium level, age, sex, baseline HAMD score) compared to a null model that contains only covariates in the broadly and narrowly defined study population.

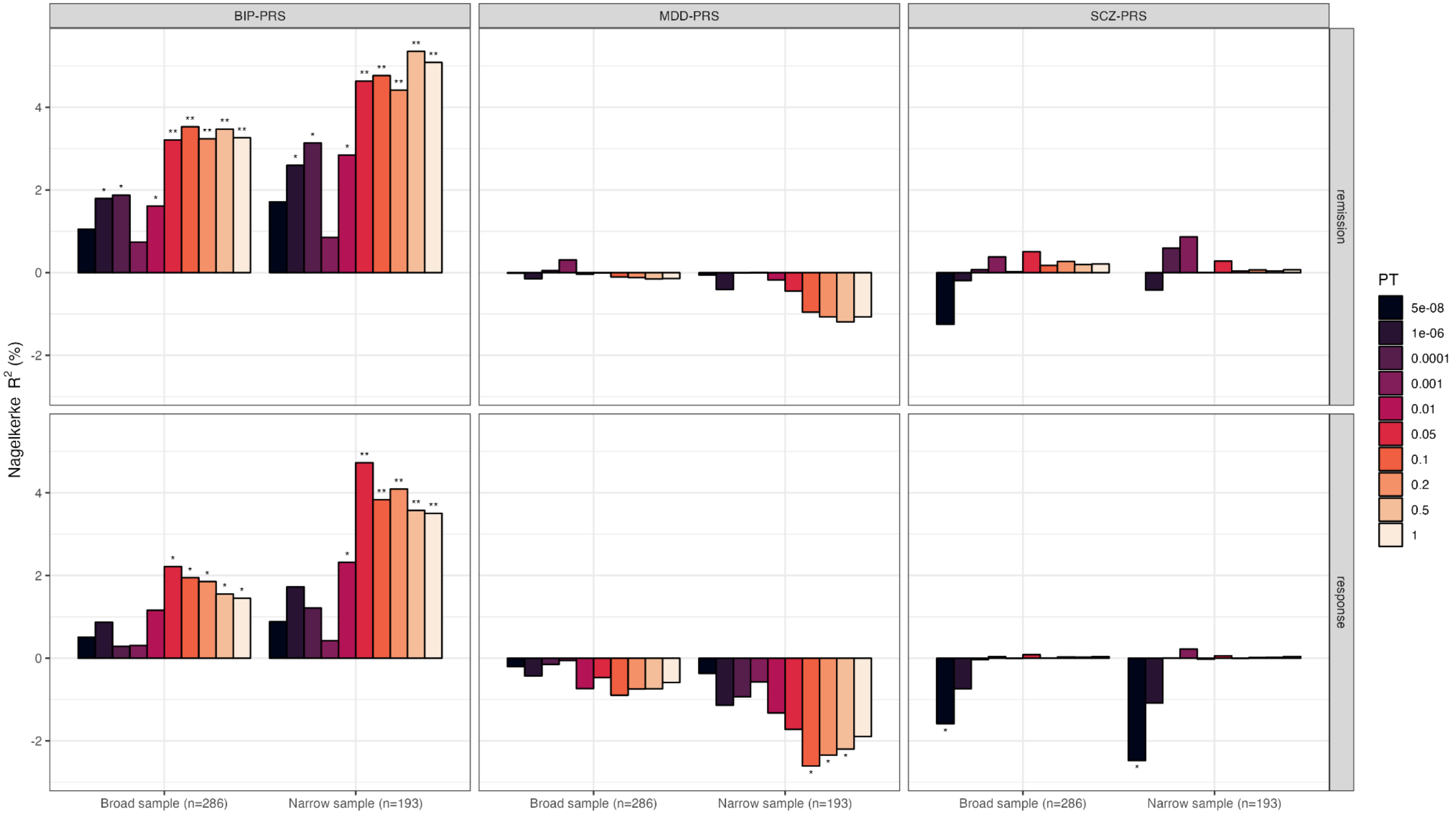


**Figure S6.** Association between polygenic risk scores for SCZ, BIP, MDD and LA outcomes (C+PT). Bar chart showing the proportion of variability (Nagelkerke $R^2$) in LA remission (upper panels) and response (lower panels) explained by BIP-, MDD- and SCZ-PRS (left, middle and right panel) in both study populations (x-axis). Negative Nagelkerke $R^2$ values reflect an inverse relationship between polygenic risk and outcome. PRS are calculated for 10 distinct p-value thresholds (PT; represented by color) using clumped summary statistics ($R^2 > 0.1$ within a 500kb window; additional filters were applied to remove SNPs with INFO < 0.9, MAF < 0.05, insertions, deletions or strand-ambiguousness). Displayed p-values adjacent to each bar: * $p < 0.05$; ** $p < 0.01$; *** $p < 0.001$.

**Table S1.** GWAS sample sizes and number of SNPs included in polygenic risk scores

| | | GWAS sample sizes | | |
|---|---|---|---|---|
| | | **BIP-PRS** (Mullins et al., 2019) | **MDD-PRS** (Adams et al., 2024) | **SCZ-PRS** (Trubetskoy et al., 2022) |
| **Sample size** | $N_{cas}$ | 41,917 | 525,006 | 67,390 |
| | $N_{con}$ | 371,549 | 3,361,321 | 94,015 |
| | $N_{eff}$ | 150,670 | 1,816,331 | 157,013 |
| | | **Number of SNPs** | | |
| **Polygenic scoring method**[a] | **PT**[b] | **BIP-PRS** | **MDD-PRS** | **SCZ-PRS** |
| PRS-CS | 1 | 886,211 | 972,563 | 983,255 |
| C+PT | 5e-08 | 62 | 788 | 284 |
| C+PT | 1e-06 | 160 | 1,400 | 549 |
| C+PT | 0.0001 | 921 | 4,204 | 2,058 |
| C+PT | 0.001 | 2,760 | 8,392 | 4,753 |
| C+PT | 0.01 | 9,458 | 19,375 | 12,775 |
| C+PT | 0.05 | 23,849 | 37,185 | 27,591 |
| C+PT | 0.1 | 35,464 | 50,289 | 38,798 |
| C+PT | 0.2 | 52,471 | 67,842 | 54,156 |
| C+PT | 0.5 | 83,209 | 97,084 | 81,356 |
| C+PT | 1 | 103,785 | 117,172 | 99,188 |

Abbreviations: LA - Lithium Augmentation, BIP - Bipolar Disorder, SCZ - Schizophrenia, MDD - Major Depressive Disorder, PRS - Polygenic risk scores, C+PT - clumping and p-value thresholding, PT - p-value threshold, $N_{cas}$ - number of cases, $N_{con}$ - number of controls, $N_{eff}$ - effective sample size of the GWAS meta-analysis

[a]PRS were obtained using two different methods: a Bayesian approach (PRS-CS) as well as a standard LD-based clumping and p-value thresholding approach (sensitivity analysis) applied to SNPs in GWAS summary statistics that overlap with LD reference panels, have a high imputation quality (INFO > 0.9) and a MAF > 0.05. PRS-CS was used with default settings (see main methods section). LD-based clumping was performed to remove variants in LD with index variants ($R^2$>0.1) within each locus (500kb window).

[b]PT denotes the upper threshold of the p-value range.

**Table S2**. Summary of antidepressants and psychotropic co-medication at baseline

| Medication | N[1] | Broad study population, N=286 | N[1] | Narrow study population, N=193 |
|---|---|---|---|---|
| Antidepressants | | | | |
| Serotonin Reuptake Inhibitor, yes (%) | 280 | 134 (47.86) | 190 | 92 (48.42) |
| Serotonin and Norepinephrine Reuptake Inhibitor, yes (%) | 279 | 83 (29.75) | 189 | 57 (30.16) |
| Tricyclic Antidepressant, yes (%) | 277 | 34 (12.27) | 188 | 21 (11.17) |
| Norepinephrine and Dopamine Reuptake Inhibitor (Bupropione), yes (%) | 277 | 14 (5.05) | 188 | 8 (4.26) |
| Valdoxan, yes (%) | 277 | 9 (3.25) | 188 | 6 (3.19) |
| Noradrenergic and specifically serotonergic antidepressant (Mirtazapine), yes (%) | 277 | 63 (22.74) | 188 | 38 (20.21) |
| Monoamine Oxidase Inhibitor (Tranylcypromine), yes (%) | 277 | 10 (3.61) | 188 | 6 (3.19) |
| Other co-medication | | | | |
| Atypical antipsychotics, yes (%) | 277 | 74 (26.71) | 188 | 50 (26.60) |
| Antiepileptic drugs, yes (%) | 277 | 25 (9.03) | 188 | 15 (7.98) |
| Benzodiazepines, yes (%) | 275 | 62 (22.55) | 186 | 43 (23.12) |
| Low-potency antipsychotics, yes (%) | 276 | 17 (6.16) | 187 | 10 (5.35) |

[1]n (%)

**Table S3.** Association between polygenic risk scores for SCZ, BIP, MDD and clinical outcomes after LA

| | | Response | | | | Remission | | | |
|---|---|---|---|---|---|---|---|---|---|
| | **PRS** | **HR (%95 CI)** | **p** | **Nagelkerke $R^2$** | **C** | **HR (%95 CI)** | **p** | **Nagelkerke $R^2$** | **C** |
| Broad study population (n=286) | | | | | | | | | |
| | BIP-PRS | 1.23 (1.02-1.48) | 2.82e-02 | 1.70 | 0.60 | 1.48 (1.18-1.86) | 8.22e-04 | 4.09 | 0.72 |
| | SCZ-PRS | 1.05 (0.87-1.27) | 5.86e-01 | 0.11 | 0.58 | 1.15 (0.91-1.45) | 2.30e-01 | 0.54 | 0.71 |
| | MDD-PRS | 0.87 (0.72-1.04) | 1.24e-01 | 0.82 | 0.58 | 0.96 (0.77-1.2) | 7.07e-01 | 0.05 | 0.70 |
| Narrow study population (n=193) | | | | | | | | | |
| | BIP-PRS | 1.29 (1.02-1.63) | 3.01e-02 | 2.50 | 0.62 | 1.52 (1.14-2.04) | 4.27e-03 | 4.53 | 0.72 |
| | SCZ-PRS | 1 (0.8-1.24) | 9.72e-01 | 0.00 | 0.59 | 1.12 (0.85-1.48) | 4.16e-01 | 0.37 | 0.71 |
| | MDD-PRS | 0.81 (0.66-1) | 4.82e-02 | 1.99 | 0.61 | 0.87 (0.68-1.12) | 2.76e-01 | 0.64 | 0.71 |

Abbreviations: LA - Lithium Augmentation, BIP - Bipolar Disorder, SCZ - Schizophrenia, MDD - Major Depressive Disorder, PRS - Polygenic risk scores, C - C-index (concordance), HR - hazard ratio, 95%CI - 95% confidence interval

**Table S4.** Hazard ratios (HR) for favorable LA outcomes by PRS strata

| | | Response | | | | Remission | | | |
|---|---|---|---|---|---|---|---|---|---|
| **PRS** | **Tertile**[b] | **NR** | **R** | **unadj. HR (%95 CI)** | **adj. HR (%95 CI)**[a] | **NRM** | **RM** | **unadj. HR (%95 CI)** | **adj. HR (%95 CI)** |
| Broad study population (n=286) | | | | | | | | | |
| MDD-PRS | 1st | 54 (56%) | 42 (44%) | 1 | 1 | 69 (72%) | 27 (28%) | 1 | 1 |
| MDD-PRS | 2nd | 49 (52%) | 46 (48%) | 1.18 (0.78-1.79) | 1.13 (0.73-1.73) | 61 (64%) | 34 (36%) | 1.27 (0.77-2.11) | 1.26 (0.74-2.13) |
| MDD-PRS | 3rd | 59 (62%) | 36 (38%) | 0.85 (0.54-1.33) | 0.84 (0.53-1.33) | 70 (74%) | 25 (26%) | 0.90 (0.52-1.55) | 0.95 (0.54-1.67) |
| BIP-PRS | 1st | 62 (65%) | 34 (35%) | 1 | 1 | 74 (77%) | 22 (23%) | 1 | 1 |
| BIP-PRS | 2nd | 53 (56%) | 42 (44%) | 1.62 (1.03-2.55) | 1.68 (1.05-2.67) | 67 (71%) | 28 (29%) | 1.60 (0.91-2.80) | 2.07 (1.16-3.69) |
| BIP-PRS | 3rd | 47 (49%) | 48 (51%) | 1.72 (1.11-2.67) | 1.75 (1.11-2.77) | 59 (62%) | 36 (38%) | 2.00 (1.17-3.40) | 2.42 (1.39-4.23) |
| SCZ-PRS | 1st | 60 (62%) | 36 (38%) | 1 | 1 | 73 (76%) | 23 (24%) | 1 | 1 |
| SCZ-PRS | 2nd | 50 (53%) | 45 (47%) | 1.19 (0.77-1.85) | 1.12 (0.72-1.75) | 60 (63%) | 35 (37%) | 1.47 (0.87-2.50) | 1.32 (0.77-2.27) |
| SCZ-PRS | 3rd | 52 (55%) | 43 (45%) | 1.31 (0.84-2.04) | 1.24 (0.79-1.96) | 67 (71%) | 28 (29%) | 1.26 (0.73-2.20) | 1.29 (0.73-2.28) |
| Narrow study population (n=193) | | | | | | | | | |
| MDD-PRS | 1st | 34 (52%) | 31 (48%) | 1 | 1 | 46 (71%) | 19 (29%) | 1 | 1 |
| MDD-PRS | 2nd | 30 (47%) | 34 (53%) | 1.20 (0.74-1.95) | 1.08 (0.65-1.80) | 38 (59%) | 26 (41%) | 1.44 (0.79-2.59) | 1.26 (0.67-2.37) |
| MDD-PRS | 3rd | 39 (61%) | 25 (39%) | 0.77 (0.46-1.31) | 0.72 (0.42-1.24) | 47 (73%) | 17 (27%) | 0.85 (0.44-1.64) | 0.86 (0.44-1.67) |
| BIP-PRS | 1st | 42 (65%) | 23 (35%) | 1 | 1 | 49 (75%) | 16 (25%) | 1 | 1 |
| BIP-PRS | 2nd | 33 (52%) | 31 (48%) | 1.71 (0.99-2.93) | 1.78 (1.02-3.10) | 45 (70%) | 19 (30%) | 1.39 (0.71-2.71) | 1.64 (0.83-3.27) |
| BIP-PRS | 3rd | 28 (44%) | 36 (56%) | 1.86 (1.10-3.15) | 2.02 (1.15-3.53) | 37 (58%) | 27 (42%) | 1.98 (1.07-3.68) | 2.26 (1.17-4.36) |
| SCZ-PRS | 1st | 36 (55%) | 29 (45%) | 1 | 1 | 47 (72%) | 18 (28%) | 1 | 1 |
| SCZ-PRS | 2nd | 36 (56%) | 28 (44%) | 0.92 (0.55-1.55) | 0.85 (0.50-1.45) | 42 (66%) | 22 (34%) | 1.23 (0.66-2.29) | 1.09 (0.57-2.07) |
| SCZ-PRS | 3rd | 31 (48%) | 33 (52%) | 1.22 (0.74-2.01) | 1.15 (0.68-1.93) | 42 (66%) | 22 (34%) | 1.26 (0.67-2.34) | 1.15 (0.59-2.22) |

Abbreviations: LA - Lithium Augmentation, BIP - Bipolar Disorder, MDD - Major Depressive Disorder, SCZ - Schizophrenia, PRS - Polygenic risk scores, NR - non-responder, R - responder, NRM - non-remitter, RM - remitter

[a]HR are adjusted for age, sex, 5 PCs, HAMD baseline score, sufficient lithium level.

[b]Strata with the lowest polygenic loadings were selected as a reference.

**Table S5.** Association between polygenic risk scores for SCZ, BIP, MDD and clinical outcomes after LA (multi-PRS model)

| | | Response | | | Remission | | |
|---|---|---|---|---|---|---|---|
| | **PRS** | **HR (%95 CI)** | **p** | **Nagelkerke $R^2$** | **HR (%95 CI)** | **p** | **Nagelkerke $R^2$** |
| Broad study population (n=286) | | | | | | | |
| | BIP-PRS | 1.31 (1.07-1.62) | 1.08e-02 | 2.26 | 1.53 (1.19-1.96) | 9.75e-04 | 3.89 |
| | SCZ-PRS | 0.99 (0.8-1.22) | 8.98e-01 | 0.01 | 1.01 (0.78-1.31) | 9.49e-01 | 0.00 |
| | MDD-PRS | 0.81 (0.66-0.98) | 2.92e-02 | 1.66 | 0.87 (0.69-1.09) | 2.25e-01 | 0.54 |
| Narrow study population (n=193) | | | | | | | |
| | BIP-PRS | 1.43 (1.1-1.86) | 6.85e-03 | 2.70 | 1.6 (1.17-2.21) | 3.76e-03 | 3.39 |
| | SCZ-PRS | 0.91 (0.7-1.17) | 4.51e-01 | 0.21 | 0.97 (0.7-1.34) | 8.43e-01 | 0.02 |
| | MDD-PRS | 0.77 (0.62-0.96) | 2.17e-02 | 1.92 | 0.82 (0.63-1.06) | 1.31e-01 | 0.92 |

Abbreviations: LA - Lithium Augmentation, BIP - Bipolar Disorder, SCZ - Schizophrenia, MDD - Major Depressive Disorder, PRS - Polygenic risk scores, HR - hazard ratio, 95%CI - 95% confidence interval

**Table S6.** Association between polygenic risk scores for SCZ, BIP, MDD at ten p-value thresholds and clinical outcomes after LA

| | | | BIP-PRS | | | MDD-PRS | | | SCZ-PRS | | |
|---|---|---|---|---|---|---|---|---|---|---|---|
| | Outcome | PT[a] | HR (95% CI)[b] | p | NKR2[c] | HR (95% CI)[b] | p | NKR2[c] | HR (95% CI)[b] | p | NKR2[c] |
| Broad study population (n=286) | | | | | | | | | | | |
| | remission | 5e-08 | 1.21 (0.97-1.52) | 9.56e-02 | 1.05 | 0.98 (0.79-1.21) | 8.44e-01 | 0.01 | 0.82 (0.66-1.01) | 6.53e-02 | 1.25 |
| | remission | 1e-06 | 1.30 (1.02-1.65) | 3.09e-02 | 1.79 | 0.93 (0.75-1.16) | 5.25e-01 | 0.15 | 0.92 (0.74-1.15) | 4.70e-01 | 0.19 |
| | remission | 0.0001 | 1.29 (1.03-1.61) | 2.43e-02 | 1.88 | 1.04 (0.84-1.30) | 7.12e-01 | 0.05 | 1.05 (0.85-1.30) | 6.60e-01 | 0.07 |
| | remission | 0.001 | 1.18 (0.94-1.47) | 1.59e-01 | 0.74 | 1.11 (0.89-1.39) | 3.60e-01 | 0.31 | 1.12 (0.90-1.39) | 3.10e-01 | 0.38 |
| | remission | 0.01 | 1.26 (1.01-1.58) | 3.90e-02 | 1.61 | 0.97 (0.78-1.20) | 7.56e-01 | 0.04 | 1.03 (0.82-1.29) | 8.17e-01 | 0.02 |
| | remission | 0.05 | 1.40 (1.12-1.74) | 3.05e-03 | 3.21 | 0.98 (0.79-1.22) | 8.83e-01 | 0.01 | 1.15 (0.91-1.44) | 2.42e-01 | 0.51 |
| | remission | 0.1 | 1.42 (1.14-1.78) | 2.14e-03 | 3.53 | 0.94 (0.76-1.17) | 5.95e-01 | 0.10 | 1.08 (0.86-1.36) | 4.92e-01 | 0.18 |
| | remission | 0.2 | 1.40 (1.12-1.76) | 3.26e-03 | 3.24 | 0.94 (0.75-1.17) | 5.71e-01 | 0.12 | 1.10 (0.88-1.38) | 3.92e-01 | 0.27 |
| | remission | 0.5 | 1.42 (1.14-1.78) | 2.11e-03 | 3.47 | 0.93 (0.74-1.16) | 5.21e-01 | 0.15 | 1.09 (0.87-1.37) | 4.65e-01 | 0.20 |
| | remission | 1 | 1.40 (1.12-1.75) | 2.83e-03 | 3.26 | 0.93 (0.75-1.16) | 5.36e-01 | 0.14 | 1.09 (0.87-1.37) | 4.50e-01 | 0.21 |
| | response | 5e-08 | 1.12 (0.93-1.34) | 2.34e-01 | 0.51 | 0.93 (0.78-1.12) | 4.48e-01 | 0.20 | 0.82 (0.69-0.98) | 3.30e-02 | 1.59 |
| | response | 1e-06 | 1.16 (0.96-1.40) | 1.20e-01 | 0.87 | 0.90 (0.75-1.08) | 2.68e-01 | 0.43 | 0.88 (0.73-1.05) | 1.46e-01 | 0.74 |
| | response | 0.0001 | 1.09 (0.91-1.30) | 3.67e-01 | 0.29 | 0.94 (0.78-1.13) | 5.16e-01 | 0.15 | 0.97 (0.82-1.16) | 7.57e-01 | 0.03 |
| | response | 0.001 | 1.09 (0.91-1.30) | 3.52e-01 | 0.31 | 0.96 (0.80-1.16) | 6.94e-01 | 0.05 | 1.03 (0.87-1.23) | 7.41e-01 | 0.04 |
| | response | 0.01 | 1.18 (0.99-1.42) | 7.18e-02 | 1.16 | 0.87 (0.72-1.05) | 1.48e-01 | 0.73 | 0.99 (0.83-1.19) | 9.46e-01 | 0.00 |
| | response | 0.05 | 1.26 (1.05-1.52) | 1.18e-02 | 2.22 | 0.90 (0.74-1.08) | 2.49e-01 | 0.47 | 1.05 (0.88-1.25) | 6.24e-01 | 0.09 |
| | response | 0.1 | 1.24 (1.04-1.49) | 1.95e-02 | 1.95 | 0.86 (0.71-1.04) | 1.12e-01 | 0.89 | 1.01 (0.85-1.21) | 9.09e-01 | 0.00 |
| | response | 0.2 | 1.23 (1.03-1.47) | 2.24e-02 | 1.86 | 0.87 (0.72-1.05) | 1.47e-01 | 0.74 | 1.03 (0.86-1.23) | 7.70e-01 | 0.03 |
| | response | 0.5 | 1.21 (1.01-1.45) | 3.57e-02 | 1.55 | 0.87 (0.72-1.05) | 1.49e-01 | 0.74 | 1.03 (0.86-1.23) | 7.78e-01 | 0.03 |
| | response | 1 | 1.20 (1.01-1.43) | 4.22e-02 | 1.45 | 0.88 (0.73-1.07) | 1.98e-01 | 0.59 | 1.03 (0.86-1.23) | 7.41e-01 | 0.04 |
| Narrow study population (n=193) | | | | | | | | | | | |
| | remission | 5e-08 | 1.27 (0.97-1.67) | 8.11e-02 | 1.72 | 0.96 (0.75-1.23) | 7.47e-01 | 0.06 | 0.74 (0.57-0.96) | 2.32e-02 | 2.80 |

| | | BIP-PRS | | | MDD-PRS | | | SCZ-PRS | | |
|---|---|---|---|---|---|---|---|---|---|---|
| Outcome | PT[a] | HR (95% CI)[b] | p | NKR2[c] | HR (95% CI)[b] | p | NKR2[c] | HR (95% CI)[b] | p | NKR2[c] |
| remission | 1e-06 | 1.36 (1.02-1.82) | 3.45e-02 | 2.60 | 0.89 (0.69-1.15) | 3.86e-01 | 0.41 | 0.89 (0.68-1.16) | 3.80e-01 | 0.42 |
| remission | 0.0001 | 1.39 (1.06-1.83) | 1.76e-02 | 3.14 | 0.98 (0.77-1.26) | 9.01e-01 | 0.01 | 1.14 (0.89-1.47) | 3.00e-01 | 0.59 |
| remission | 0.001 | 1.19 (0.90-1.57) | 2.15e-01 | 0.85 | 1.01 (0.78-1.31) | 9.53e-01 | 0.00 | 1.17 (0.92-1.50) | 2.08e-01 | 0.87 |
| remission | 0.01 | 1.36 (1.04-1.77) | 2.40e-02 | 2.85 | 0.93 (0.72-1.20) | 5.69e-01 | 0.18 | 1.01 (0.79-1.30) | 9.20e-01 | 0.01 |
| remission | 0.05 | 1.48 (1.14-1.93) | 3.61e-03 | 4.63 | 0.89 (0.69-1.15) | 3.65e-01 | 0.45 | 1.10 (0.85-1.43) | 4.74e-01 | 0.28 |
| remission | 0.1 | 1.48 (1.14-1.93) | 3.46e-03 | 4.77 | 0.84 (0.65-1.09) | 1.86e-01 | 0.96 | 1.04 (0.80-1.34) | 7.87e-01 | 0.04 |
| remission | 0.2 | 1.46 (1.12-1.90) | 4.75e-03 | 4.42 | 0.83 (0.63-1.08) | 1.64e-01 | 1.07 | 1.05 (0.81-1.36) | 7.26e-01 | 0.07 |
| remission | 0.5 | 1.52 (1.17-1.96) | 1.65e-03 | 5.35 | 0.82 (0.62-1.07) | 1.41e-01 | 1.19 | 1.04 (0.80-1.35) | 7.86e-01 | 0.04 |
| remission | 1 | 1.49 (1.16-1.93) | 2.10e-03 | 5.08 | 0.83 (0.63-1.08) | 1.63e-01 | 1.07 | 1.05 (0.81-1.36) | 7.22e-01 | 0.07 |
| response | 5e-08 | 1.16 (0.93-1.44) | 2.00e-01 | 0.88 | 0.92 (0.75-1.12) | 4.01e-01 | 0.37 | 0.78 (0.63-0.98) | 2.89e-02 | 2.47 |
| response | 1e-06 | 1.23 (0.98-1.54) | 7.42e-02 | 1.72 | 0.85 (0.69-1.05) | 1.39e-01 | 1.14 | 0.85 (0.69-1.06) | 1.48e-01 | 1.08 |
| response | 0.0001 | 1.18 (0.95-1.46) | 1.29e-01 | 1.21 | 0.87 (0.71-1.07) | 1.80e-01 | 0.93 | 1.01 (0.83-1.24) | 9.15e-01 | 0.01 |
| response | 0.001 | 1.11 (0.89-1.38) | 3.69e-01 | 0.43 | 0.89 (0.72-1.11) | 2.94e-01 | 0.58 | 1.07 (0.88-1.30) | 5.16e-01 | 0.22 |
| response | 0.01 | 1.26 (1.01-1.56) | 3.68e-02 | 2.32 | 0.83 (0.67-1.04) | 1.10e-01 | 1.32 | 0.98 (0.80-1.19) | 8.36e-01 | 0.02 |
| response | 0.05 | 1.41 (1.13-1.76) | 2.66e-03 | 4.73 | 0.81 (0.65-1.02) | 6.88e-02 | 1.72 | 1.04 (0.85-1.27) | 7.36e-01 | 0.06 |
| response | 0.1 | 1.35 (1.08-1.67) | 7.30e-03 | 3.84 | 0.77 (0.62-0.97) | 2.58e-02 | 2.61 | 1.00 (0.82-1.22) | 9.91e-01 | 0.00 |
| response | 0.2 | 1.35 (1.09-1.67) | 5.48e-03 | 4.09 | 0.78 (0.62-0.98) | 3.54e-02 | 2.34 | 1.02 (0.83-1.25) | 8.59e-01 | 0.02 |
| response | 0.5 | 1.32 (1.07-1.63) | 8.83e-03 | 3.58 | 0.79 (0.63-0.99) | 4.13e-02 | 2.20 | 1.02 (0.83-1.26) | 8.28e-01 | 0.02 |
| response | 1 | 1.32 (1.07-1.62) | 9.48e-03 | 3.51 | 0.80 (0.64-1.01) | 5.84e-02 | 1.89 | 1.03 (0.84-1.27) | 7.77e-01 | 0.04 |

Abbreviations: LA - Lithium Augmentation, BIP - Bipolar Disorder, SCZ - Schizophrenia, MDD - Major Depressive Disorder, PRS - Polygenic risk scores, HR - hazard ratio, 95% CI - 95% confidence interval, NKR2 - Nagelkerke $R^2$

[a]To obtain LD-independent variants in each GWAS training data set, we performed clumping ($R^2 > 0.1$ within a 500kb window) on all SNPs that overlap with the LD reference panel (European subset of the HRC), have a MAF > 0.05 and good imputation quality (INFO scores > 0.9), are not strand-ambiguous, insertions or deletions. PRS were then calculated for a range of p-value thresholds (PT) as described in the main method section.

[c]Nagelkerke $R^2$ (%) represents the amount of variation in response and remission that is attributable to each PRS.

[b]HR > 1 indicates a favorable outcome with increasing polygenic risk.
